# Focal cerebral arteriopathy in adults: A single centre experience

**DOI:** 10.1101/2025.10.03.25337301

**Authors:** Deepak Menon, Thuppanattumadam Ananthasubramanian Sangeeth, Subasree Ramakrishnan, Manisha Gupta, Karthik Kulanthaivelu, Allen Johnson, Pritam Raja, Jitender Saini, Girish B. Kulkarni, P. R. Srijithesh

## Abstract

**Background and Objectives:** Focal cerebral arteriopathy (FCA) is a recognized cause of pediatric stroke, but its presentation in adults is poorly defined, with limited cohorts and scarce advanced imaging data. We aimed to describe the clinical spectrum, radiological features, treatment strategies, and outcomes of adult FCA in the largest single-center cohort to date.

**Methods:** We retrospectively reviewed consecutive adults (>18 years) with ischemic stroke admitted between 2017 and 2024. FCA was defined as unilateral focal stenosis/irregularity of anterior circulation arteries (terminal ICA, M1, M2, A1) after excluding mimics such as vasculitis, dissection, embolic occlusion, and intracranial atherosclerosis. All underwent MRI with contrast-enhanced vessel wall imaging and confirmation on a second modality. Clinical, radiological, and outcome data were collected. Severity was scored using the FCA Severity Score (FCASS).

**Results:** Of 2,237 stroke admissions, 47 patients (2.1%) met criteria for FCA. Median age was 30 years, with near-equal sex distribution. Hemiparesis with or without aphasia predominated, and strokes were generally mild (median NIHSS 3, mRS 1.2). Preceding clustered TIAs occurred in one-third, and 32% reported new unilateral headache. Infarcts often involved lenticulostriate and MCA watershed territories. Median FCASS was 4. Vessel wall imaging showed concentric enhancement in 74.5%. All patients received antiplatelets; 49% received additional immunosuppression, most often IV methylprednisolone. Over a median 6-month follow-up (mean 12.4), no stroke recurrences occurred, though 5 patients showed radiological progression and 2 developed contralateral stenosis. Functional outcomes were favorable, with 87% achieving mRS 0-2.

**Discussion:** Adult FCA is uncommon but clinically distinct, marked by mild strokes, clustered TIAs, and concentric vessel wall enhancement. The course was largely monophasic, with favorable functional outcomes and no recurrent strokes, regardless of immunosuppressive therapy. Radiological progression was rare but included contralateral involvement, raising the possibility of overlap with unilateral moyamoya disease. Vessel wall imaging is valuable for diagnosis, and longer follow-up is needed to clarify pathogenesis and treatment strategies.

## Introduction

Focal cerebral arteriopathy (FCA) is a well-established etiology of ischemic stroke in the pediatric population, characterized by unilateral, localized stenosis of large intracranial arteries, primarily within the anterior circulation, resulting in focal cerebral infarctions.[1] FCA can superficially resemble other intracranial vasculopathies such as Moyamoya disease (MMD), intracranial atherosclerosis (ICAD), intracranial arterial dissection, embolic occlusion and central nervous system (CNS) vasculitis but has distinct clinical and radiological features.[2] Although its precise pathophysiological mechanism remains uncertain, current evidence suggests a post-infectious or inflammatory vasculopathy as a potential underlying process.[3]

Although FCA is well-recognized as a cause for pediatric stroke, it appears to be less prevalent than initially suspected, and remains rare in in adults.[4] The largest single-center adult cohort to date comprises of fourteen patients, in whom a generally benign clinical course was observed.[5] However, the imaging modalities employed in that study were limited to MR or CT angiography. Advanced imaging techniques, such as contrast-enhanced MR vessel wall imaging which is known to provide superior specificity in differentiating FCA from other vasculopathies, were not utilized.

Moreover, the optimal therapeutic approach to FCA in adults remains unclear. While corticosteroids have been explored and is generally recommended in pediatric cases given the presumed inflammatory basis, their efficacy remains inconclusive in adults[6,7] Data regarding the clinical spectrum, radiologic characteristics, treatment strategies, and long-term outcomes of FCA in adults are thus scarce. In this context, we present the largest, single-center cohort of adults with FCA-associated stroke, with the clinicodemographic pattern, neuroimaging findings, therapeutic interventions, and longitudinal follow-up.

## Methods

We adopted the Wintermark’s definition of FCA which defines focal and unilateral stenosis/irregularity of the large intracranial arteries of the anterior circulation distal internal carotid artery and its proximal branches namely terminal ICA, M1, M2 and A1 where no alternate causes could be found.[1] To exclude alternate causes such as CNS vasculitis, intracranial dissection, embolic occlusion and intracranial atherosclerotic disease we used the following exclusion criteria: patients with calcification or lipid-laden intracranial stenosis and extracranial atherosclerotic changes (ICAD), hematoma in vessel wall or pseudolumen and/ or a history of head trauma (intracranial dissection), abrupt occlusion of lumen with presence of intraluminal thrombus (embolic occlusion) or having other features of CNS vasculitis including multiple territory infarcts, parenchymal microbleeds or multiple vessel involvement.[5,8–10] We also excluded any patient who had an alternate etiology including cardiac abnormalities, inflammatory markers or a known history of fever and rash suggestive of varicella zoster or herpes zoster infection in the preceding one year.

Data from consecutive patients admitted and evaluated in the stroke unit from 2017 to 2024 were collected from chart review. Patients who had a minimum of 3 months of clinical and/or radiological follow-up were included. All patients who were included had the vessel findings verified through 2 different modalities-either MRI, CTA or DSA. All patients had at least one MRI with contrast enhanced vessel wall imaging.

Patient clinical data, including stroke syndrome, severity, neuroimaging findings, clinical course, and outcome were reviewed. The FCA Severity Score (FCASS) was determined for each angiogram as previously described using the following 4-point scale: no involvement (0), irregularity or banding without stenosis (1), <50% stenosis (2), >50% stenosis (3), and occlusion (4).[11] The 5 arterial segments assessed were the supraclinoid internal carotid artery, A1, A2, M1, and M2. Any degree of stenosis other than occlusion involving the A2 and M2 branches was scored as 3. Total FCASS was the sum of arteriopathy severity for each segment ranging from 0 to 20. Stroke progression was defined as worsening modified Alberta Stroke Program Early CT Score (mod ASPECTS) on follow-up neuroimaging within 4 weeks of the initial stroke presentation, indicating infarct in new brain regions. Stroke recurrence was defined as a new, acute infarct on MRI diffusion weighted imaging, more than 4 weeks from the initial stroke presentation and after the patient had returned to the new clinical baseline. Radiological progression was defined if patients had new infarcts, progression of focal stenosis or increase in vessel wall contrast enhancement in follow-up images.

Patients underwent the stroke protocol MRI including all conventional two dimensional 2D MRI sequences as T2W, T1W, FLAIR (fluid attenuation inversion recovery), SWI (susceptibility weighted Imaging), DWI (diffusion weighted imaging), Time of Flight MR angiogram, contrast angiogram of the neck vessels and post contrast 3D T1 weighted black blood vessel wall imaging(VWI)-Siemens VIDA VISTA-(64 channel coil, SPIR sequence 1mm slice thickness with 0.5mm slice gap, TR/TE-600/28 ms, 250 x 250 matrix). All the MRIs were reviewed by neuroradiologists KK and AL and any dispute settled by JS who is the senior interventional neurologist with more than 20 years of experience.

Data were analyzed using IBM SPSS Statistics version 24. Continuous variables were summarized as mean ± standard deviation (SD) or median with interquartile range (IQR), depending on distribution. Categorical variables were expressed as frequencies and percentages. Between-group comparisons (e.g., patients receiving immunosuppressive therapy vs. those who did not) were performed using the Chi-square test or Fisher’s exact test for categorical variables, and the independent samples *t*-test or Mann–Whitney *U* test for continuous variables, as appropriate. A *p*-value <0.05 was considered statistically significant. Due to the limited number of outcome events, multivariable regression analysis was not performed.

## Standard Protocol Approvals, Registrations, and Patient Consents

This study was approved by the Institutional Ethics Committee of NIMHANS, Bangalore, India. The requirement for written informed consent was waived due to the retrospective nature of the study.

## Results

During the study period from June 2017 to December 2024, a total of 2,237 patients aged over 18 years were admitted to the stroke care services (ICU and wards) with acute or subacute ischemic strokes. Among this cohort, 47 patients met our selection criteria for adult FCA, yielding a prevalence of 2.1%. The study work flow is provided in figure 1. The median age was 30 years and the gender distribution was essentially equal (24 females, 23 males). Although a preceding vascular event was not found in majority, 1/3^rd^ of the patients (16, 34%) has a preceding TIA or stroke before the presenting event. Preceding new onset unilateral headache was reported by 15 (31.9%) patients and one patient was 25 days postpartum and another patient was receiving anti-rabies vaccination. The median number of preceding TIAs were 3, ranging up to 10. A clear referral diagnosis was available for 32 patients which included vasculitis (12, 37.5%), FCA (7, 21.9%), MMD (5, 15.6%), ICAD (3, 9.4%), lacunar stroke (2, 6.3%), varicella vasculopathy (1, 3.1%), embolic stroke (1, 3.1%) and demyelination (1, 3.1%).

**Figure 1:**
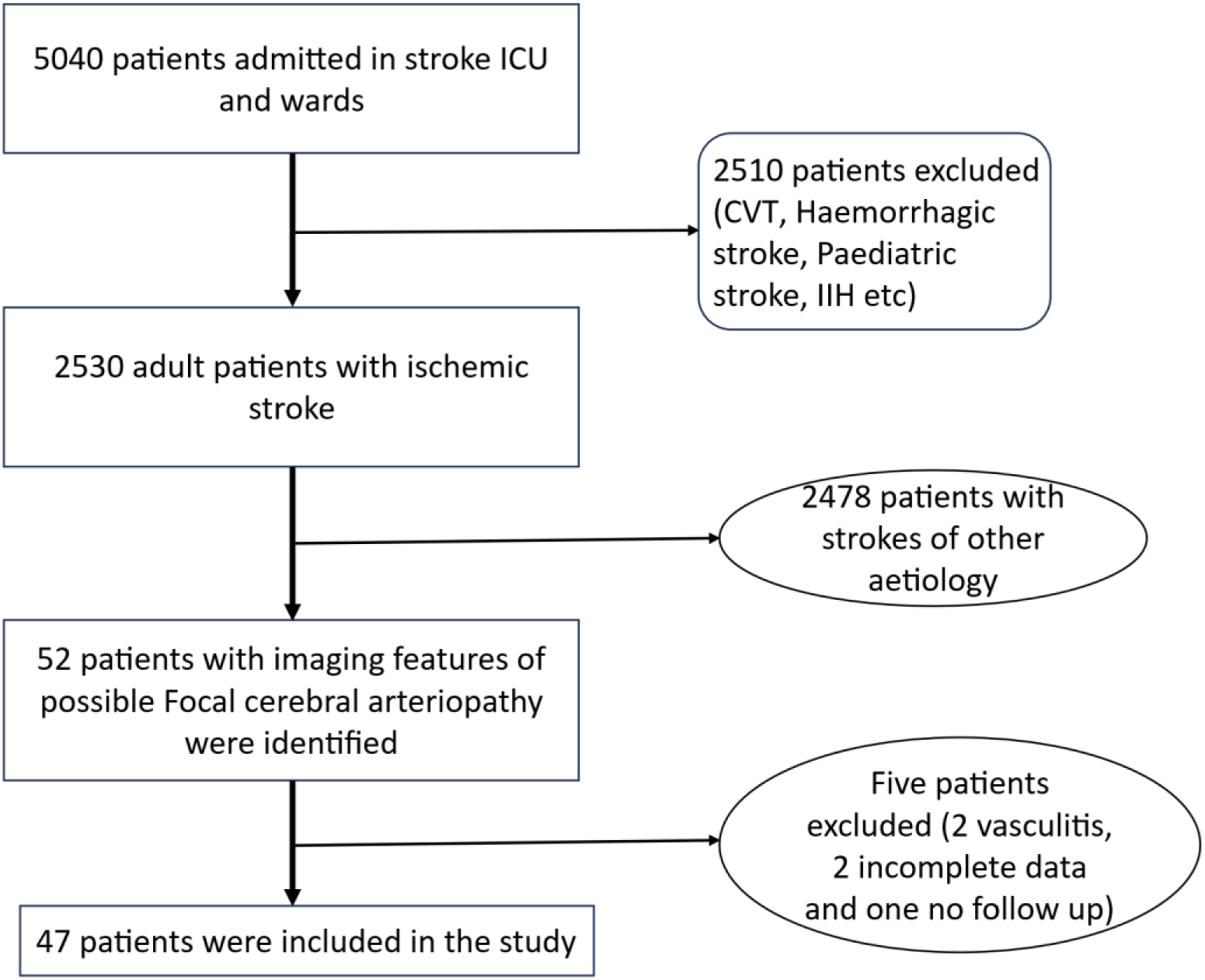
Flow chart showing the study protocol

The most common presenting syndrome were hemiparesis with or without aphasia followed by TIA and all patients had a minor severity of stroke with mean NIHSS and mRS of 3 and 1.2 respectively. Thirty-five (74.4%) of patients had an NIHSS between 0-5, 11 (23.4%) between 6-15 and 1 patient had an NIHSS of 18 at presentation. Majority of patients (35, 74.4%) had an mRS of 0-2 and 12 (25.5%) had an mRS of 3-4. The most common locations of infarcts were in the lenticulostriate territories and the MCA watershed territories as detailed in table 1, with 5 patients having no infarcts. At presentation the focal stenosis was most commonly seen in M1, terminal ICA, A1 and M2 in that order with a median FCASS sum score of 4 (mean 4.8, 2-9).

**Table 1:**
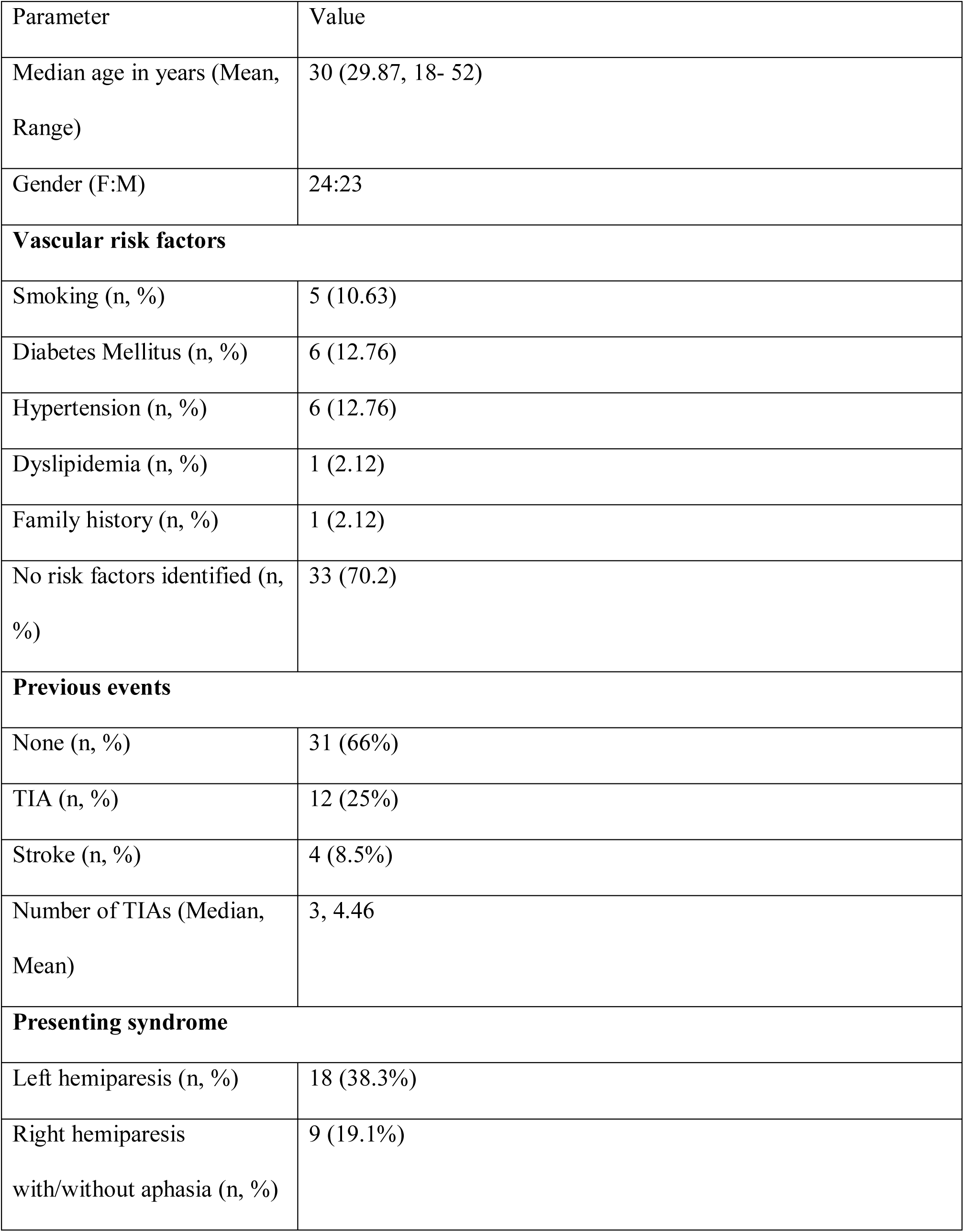
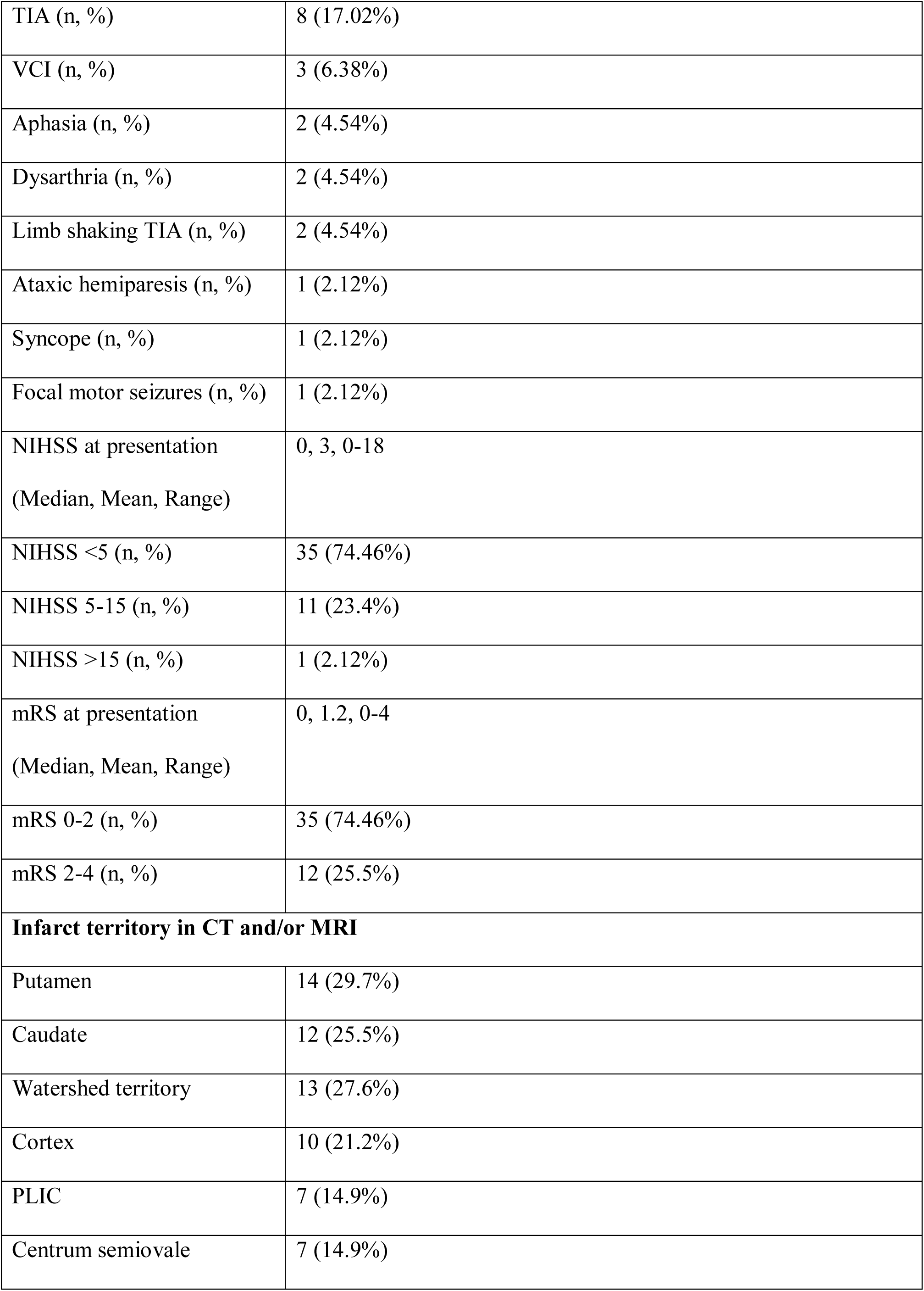
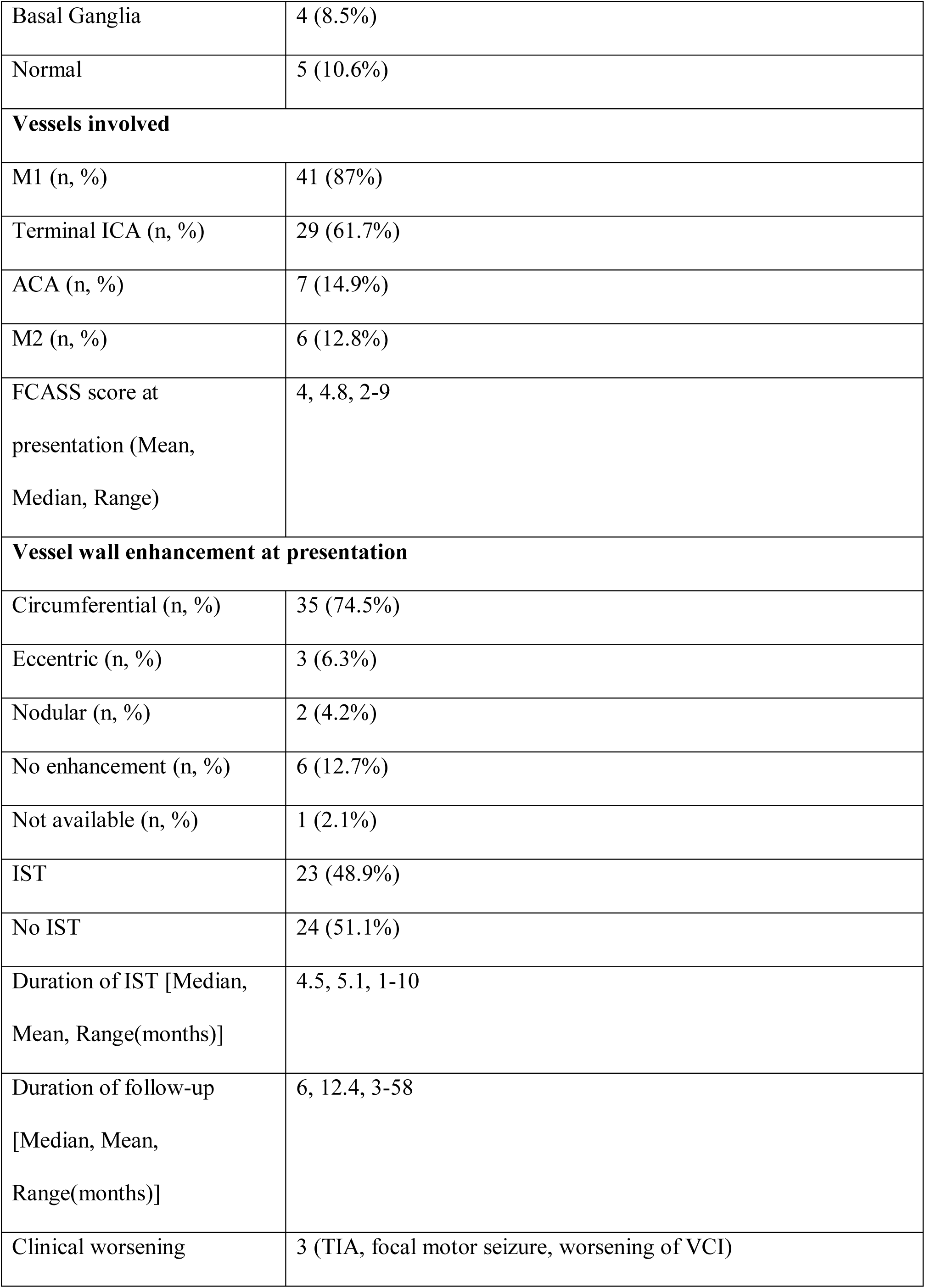
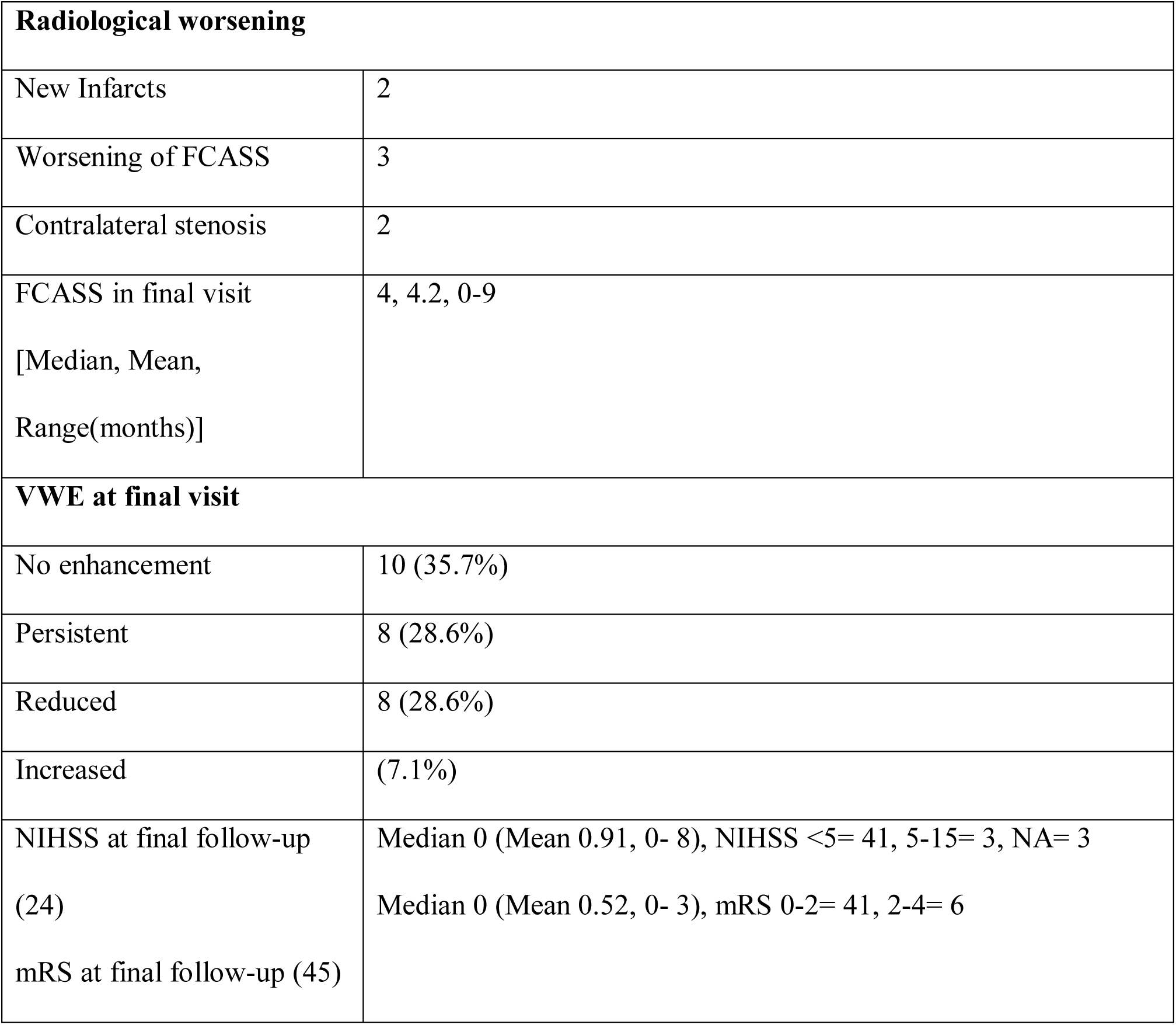
Table demonstrating the clinical, radiological and follow-up details of the patients with Focal cerebral arteriopathy from our cohort

Of the MR vessel wall imaging available (46/47 patients) a circumferential enhancement was found in 35 (74.5%), eccentric 3 (6.3%), nodular 2 (4.2%) and 6 (12.7%) had no enhancement (Figure 2). All patients received antiplatelets, with 17 (36.2%) receiving single antiplatelet and statin, 27 (57.4%) patients receiving dual antiplatelets and statins for 21 days followed by single antiplatelets and statins while 3 (6.3%) receiving only single antiplatelet. Immunosuppressive therapy was received by 23 (48.9%) of patients with a median duration of 4 months (mean 5.1, 1-10 months). The most common protocol being monthly 5 days pulse of IVMP. Other regimens included oral steroids and steroid sparing agents including AZA or MMF. Patients who received IST did not differ significantly from those who did not in terms of age, gender, preceding events, baseline NIHSS or mRS scores, initial CT ASPECTS, or vessel wall imaging findings. The mean duration of followup of patients on IST was 13.4± 9.8 months while that of no IST was 11.4±15.0 months with no significant difference (t 0.531, p 0.216). Clinical exome sequencing was performed in four of the patients and all were non-contributory.

**Figure 2.**
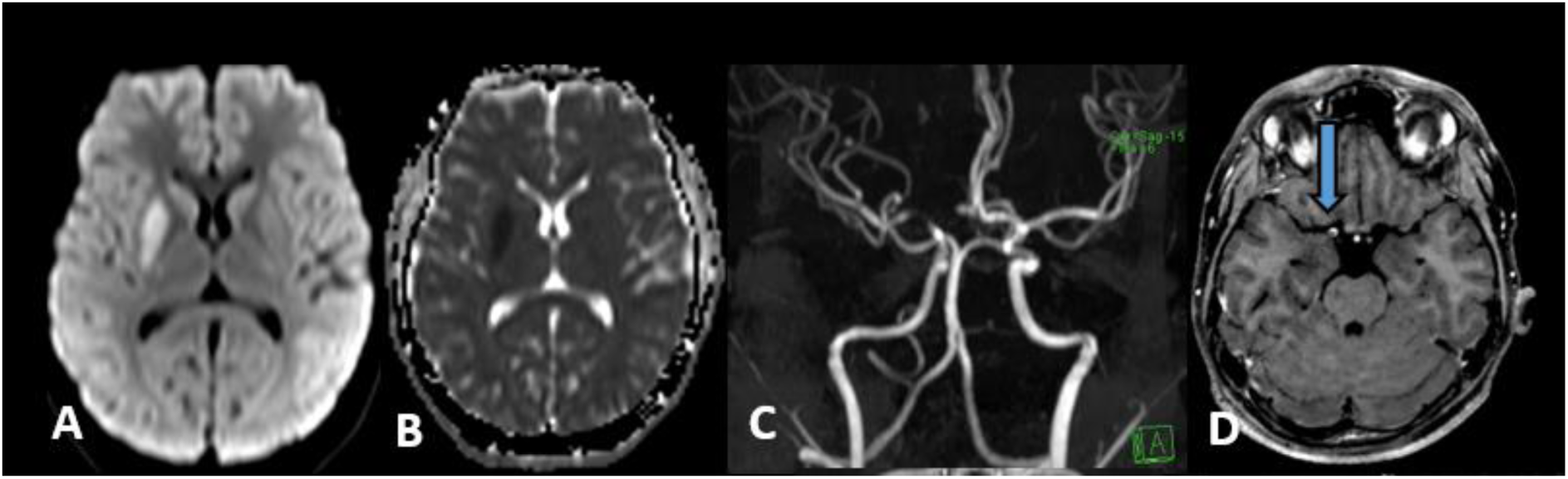
Classic case of FCA (Patient 17) (A) Axial DWI MRI image showing hyperintensity in the right putamen with (B) Corresponding hypointensity in ADC map, indicating an acute infarct; (C) TOF MR angiography of the same patient demonstrating significant narrowing of the proximal right MCA with (D) Concentric vessel wall enhancement on post contrast T1 weighted vessel wall imaging (Blue arrow)

The median follow-up period was 6 months (mean 12.4 months, 3-58 months) and 18 patients had a minimum follow-up of 12 months (mean 24.6, 6-58 months). During the follow-up, stroke progression or stroke recurrence were not found in any patient but 3 patients had clinical events in the form of one TIA, focal motor seizures with preserved awareness and mild worsening of vascular cognitive impairment. Three or more follow-up images (MRI with MRA) were available for 29 patients, 2 follow-up images were available for 12 patients and 6 patients had one follow up image. The median interval from 1^st^ to final image was 3 months (Mean 5.8 months, 3 to 58 months). Radiological worsening was found in 5 patients with new infarcts in 2 patients, worsening of FCASS score in 3 and involvement of contralateral vessels without collaterals in 2. There was a mild reduction in mean FCASS sum score at the final follow-up (4.8 to 4.2). Among the 28 patients who had a follow-up vessel wall imaging at the final follow-up, majority had either reduction (Figure 3) or resolution of vessel wall enhancement (18, 64.3%), 8 (28.6%) had persistent enhancement and only 2 (7.1%) had a worsening (Figure 4), At the last follow-up the majority of patients had improvement in good functional outcome with an mRS of 0-2 in 41 (87.2%) and NIHSS score of 0-5 in 41 (93.1%). As stroke progression or recurrence was not found in any patient and as only a very few number of patients had radiological worsening, a regression analysis for outcome predictors was not performed.

**Figure 3.**
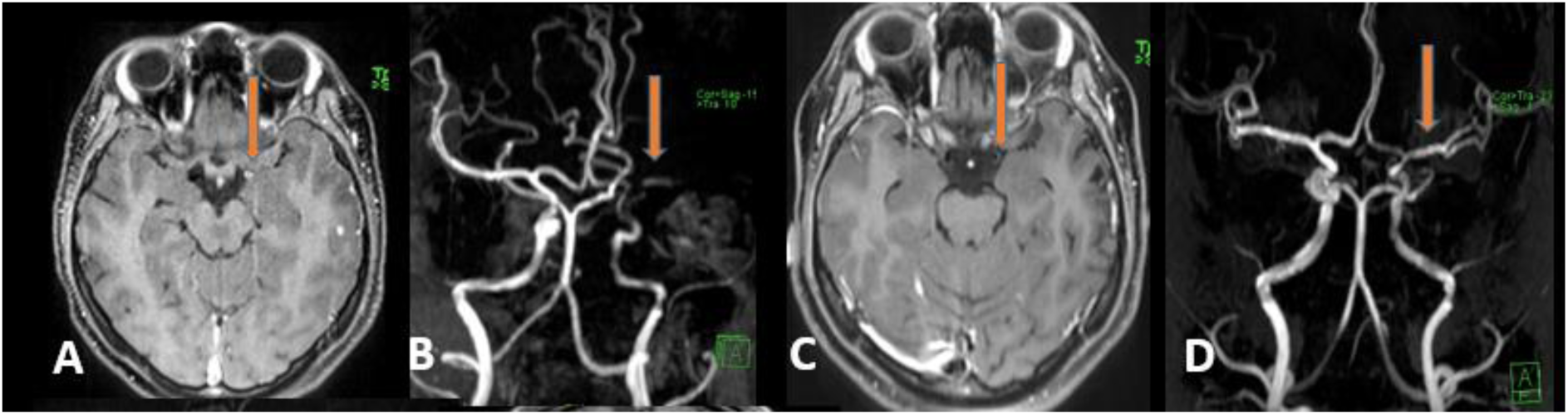
Case of FCA with resolution on follow up (Patient 20) (A) T1 weighted post contrast vessel wall imaging showing enhancement of left MCA (Orange arrow) with (B) significant luminal narrowing on TOF-MRA (Orange arrow); (C) Follow up imaging of the same patient shows reduction in vessel wall enhancement (Orange arrow) with (D) resolution of luminal narrowing on TOF-MRA (Orange arrow)

**Figure 4.**
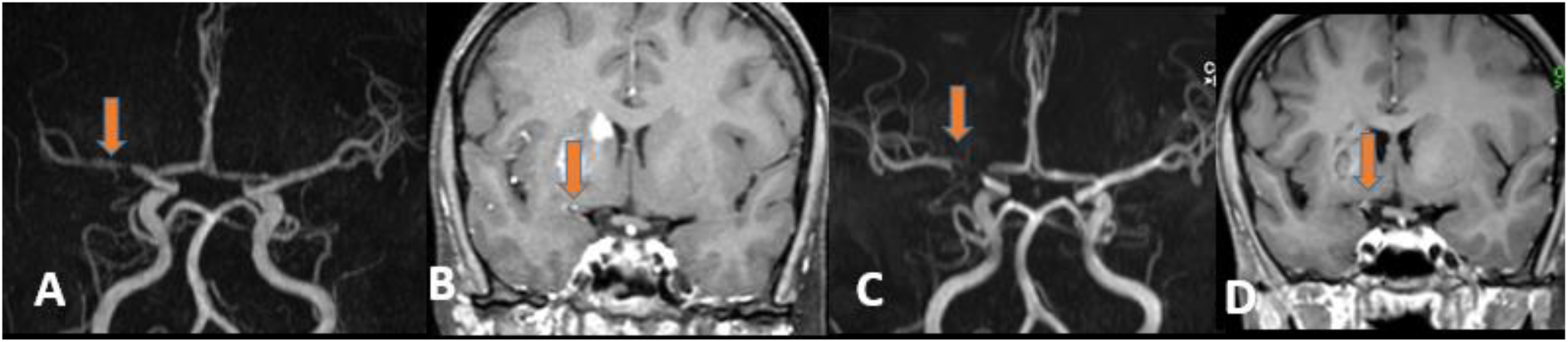
Case of FCA with progression of narrowing (Patient 41) (A) TOF-MRA showing significant luminal narrowing of right proximal MCA (Orange arrow) with (B) Vessel wall enhancement (Orange arrow); (C) Follow up MRA-TOF of P-41 demonstrating progression of luminal narrowing (Orange arrow) with (D) Decrease in vessel wall enhancement (Orange arrow)

## Discussion

The current study in adult patients with FCA, after excluding alternative etiologies such as dissection and other inflammatory arteriopathies, highlights certain distinctive clinical and radiological characteristics. The findings reaffirm the largely benign and monophasic course of FCA in majority of adults with no observed stroke recurrences and only a minority demonstrating radiological progression. Notable diagnostic clues include a preceding cluster of TIAs, not infrequent reports of new-onset unilateral headache, and the high prevalence of concentric vessel wall enhancement on MRI; the latter can be crucial for differentiating from ICAD or dissection. The infarcts are equally likely to involve the watershed territories, in contrast to the predominantly lenticulostriate involvement observed in children. The utility of steroids and other immunosuppressive therapies remains uncertain, as improvements in FCASS scores, vessel wall imaging, as well as the absence of clinical recurrences, were observed irrespective of treatment. Intriguingly, contralateral vessel narrowing without collateral formation was identified in two patients, raising the possibility that FCA may, in a subset of cases, represent an early or transitional phase in the pathogenesis of moyamoya disease but would require longer follow-up.

In a recent systematic review and meta-analysis the prevalence of FCA was reported to be 16.6% in children, lower compared to with findings from focused pediatric FCA cohorts by Oesche et al.[4,12]. In adults, although data is limited, the prevalence of FCA is much less with 2.1% in the current study and around 4-5% in other studies from western population.[5,13] But FCA in pediatric population has been traditionally divided into an FCA-inflammation type, FCA-dissection type with intracranial dissection and FCA-undetermined type.[1] However, grouping intracranial vasculopathies such as dissection and, in particular, intracranial atherosclerotic disease (ICAD), the latter being notably prevalent among adults in South Asian populations, under FCA may not be appropriate, given their distinct pathophysiological mechanisms and clinical trajectories and hence was excluded from the current cohort.[2,14] A preceding vascular event or TIA was absent in the majority of patients in the current cohort. However, when present, patients often experienced a cluster of TIAs, sometimes exceeding ten episodes, and nearly one third reported a preceding unilateral headache. Both features are uncommon in previously published adult FCA series.[5,13,15] This variation may reflect differences in the timing of patient presentation, as the absence of stroke progression or recurrence in any patient, each of whom received antiplatelet therapy upon admission, suggests that early presentation and prompt initiation of secondary prevention could play a protective role. This observation, combined with the frequent involvement of watershed and cortical territories, in addition to the lenticulostriate regions suggesting a hemodynamic component, underscoring the complex and still uncertain pathophysiological mechanisms underlying FCA. The concentric vessel wall enhancement which was seen in most of our patients has been previously reported in some pediatric and adult series of FCA and has been suggested as a criterion of steroid treatment.[3,15–17] Inflammation could be cause of the concentric vessel enhancement, edema and the ensuing stenosis resulting in infarct and hence the use of steroid would be logical backed up by initial clinical evidence.[6] However the current study failed to show any definite benefit and the result of the FOCAS trial in children examining if early steroids prevent progression is awaited.[18] Further insights into the pathogenesis may require experimental animal models or pathological validation through tissue-based studies, but would be difficult to obtain.

During follow-up, a minority of patients demonstrated clinically silent radiological progression, including new infarcts, an increase in FCASS scores, and early involvement of contralateral vessels in the absence of collateral formation. These findings raise several important questions. Unilateral moyamoya disease (U-MMD) is a key differential diagnosis for FCA, with the presence of collateral vessels often serving as the only distinguishing feature, rendering differentiation at initial presentation challenging. Notably, vessel wall enhancement may also be seen in MMD and can vary depending on age, arterial segment involved, and disease stage.[19] This raises the possibility that a subset of FCA cases may represent a transitional phase toward MMD.[20] Given that FCA is presumed to be primarily inflammatory in nature, whereas MMD is thought to be an idiopathic steno-occlusive arteriopathy with potential genetic influence, the role of CSF and serum inflammatory markers, as well as broader implementation of genetic screening for RNF213, warrants further exploration. Although FCA is typically considered a benign, monophasic, and non-progressive disorder, these observations underscore the importance of follow-up imaging to monitor for disease progression. Extended longitudinal studies in adult FCA cohorts are essential to better define its natural history.

Our study had a few limitations given its retrospective nature. The follow-up duration was relatively short and heterogenous with less than half the patients having a follow up beyond 12 months duration was relatively short restricts the ability to assess delayed disease progression, or recurrence of vascular events. Initiation of steroids and IST was heterogenous and depended on the physician’s discretion and revealed that IST was initiated for patients with more severe stroke and hence this could have influenced the apparent lack of benefit. As CSF PCR analysis was not done in all patients a varicella zoster related FCA cannot be excluded however none of the patients in the cohort had historically had varicella or herpes zoster preceding the onset of symptoms.

Extended longitudinal clinical and radiological follow-up of adult FCA cohorts is essential to better characterize its natural history, particularly given that a subset may exhibit overlapping features with unilateral moyamoya disease (U-MMD) and warrant closer surveillance. Future studies integrating inflammatory biomarkers and genetic profiling may offer deeper insights into the underlying pathophysiology and potential disease modifiers of FCA.

## Data Availability

The datasets generated and/or analyzed during the current study are not publicly available due to patient privacy and institutional regulations but are available from the corresponding author on reasonable request.

## Acknowledgements

None

